# Beliefs, Perceptions, and Behaviors Regarding Chronic Lung Disease in Kyrgyzstan: A Mixed-method FRESHAIR Study

**DOI:** 10.1101/2025.11.19.25338955

**Authors:** Chao Sun, Marise J. Kasteleyn, Evelyn A. Brakema, Charlotte C. Poot, Rianne M.J.J van der Kleij, Niels H. Chavannes, Maamed Mademilov, Talant Sooronbaev, Eline Meijer

**Author notes:** Corresponding author: Chao Sun, MPH Department of Public Health and Primary Care, Leiden University Medical Centre Hippocratespad 21, Leiden, The Netherlands.

## Abstract

**Introduction:** Chronic lung disease (CLD) represents a substantial health burden in low- and middle-income countries (LMICs). However, in-depth CLD studies in Kyrgyzstan, where CLD is highly prevalent, remain scarce.

**Objectives:** To explore in Kyrgyzstan (1) beliefs, perceptions and behaviors regarding CLD among community members (CMs), (2) relations between illness perceptions, demographics, behaviors and setting (highlands vs. lowlands), and (3) illness perception profiles across subgroups.

**Methods:** Qualitative data were gathered through semi-structured interviews and focus groups and analyzed using a Framework Method. Quantitative data were collected via questionnaires including the brief Illness Perception Questionnaire (B-IPQ), measures of smoking, indoor heating and cooking, and help-seeking behaviors. Univariable and multivariable linear regression analysis and latent class analysis (LCA) were conducted. Triangulation integrated qualitative and quantitative findings.

**Results:** We included 8 interviews, 13 focus groups, and 420 questionnaires. CMs held misperception about CLD and its causes, continued smoking despite perceived harms and prevention efforts, used risk fuels with limited ventilation, and delayed help-seeking. CMs obtained a mean B-IPQ score of 23.4 (SD 2.1), with longer duration of education, lowland setting, and being unemployed significantly associated with higher B-IPQ scores (*p*<0.05). LCA identified 2 subgroups: highlanders were more likely to be in class 1 whereas lowlanders in class 2 (χ2=5.95, *p*=0.01).

**Conclusion:** Misbeliefs and misperceptions about CLD, smoking, household risk behaviors, and delayed help-seeking were prevalent in Kyrgyz CMs. Longer education, being unemployed and living in the lowlands were associated with more negative illness perceptions. Two distinct subgroups based on B-IPQ profiles were identified: one with more negative perceptions in the consequence, symptoms, concern and emotion, while the other perceived more negatively in the timeline, treatment and understanding. Targeted education in CLD, smoking cessation support should be provided for the public to improve the awareness, prevention, and care of CLD in Kyrgyzstan.

**Summary box:** *What is already known in this topic:* CLD poses a large burden in Kyrgyzstan and other LMICs. Interventions to address non-communicable diseases in Kyrgyzstan appear to be ineffective.

**What this study adds:**

- Misconceptions about CLD, prevalent smoking, household risk behaviors, delayed help-seeking were evident in Kyrgyzstan, shaped by socioeconomic, cultural and policy-related factors.
- Infrastructure issues, affordability, accessibility to clean energy and healthcare service, weak enforcement for tobacco control and antibiotics use were barriers to CLD prevention and care.
- Longer education, being unemployed and living in the lowlands were associated with more negative illness perceptions, with distinct B-IPQ profiles in different subgroups.

**How this study might affect research, practice or policy:**

- Future studies should explore the social and cultural factors contributing to CLD misperceptions in depth and assess the effectiveness of educational interventions to improve disease understanding and help-seeking.
- Behavior change techniques should be integrated into tailored health interventions to ensure better effectiveness in improving CLD awareness and promoting behavioral change.
- Policymakers should prioritize smoking cessation programs, enforce stronger tobacco control measures, and invest in clean energy infrastructure and equitable healthcare access.

## Introduction

Chronic lung disease (CLD) encompasses a group of long-term conditions affecting the airways and lungs, including chronic obstructive pulmonary disease (COPD), asthma, pneumonitis, and other lung conditions^1^. Main risk factors for CLD include direct factors, such as smoking, indoor and outdoor air pollution and occupational exposure, as well as indirect factors like low socioeconomic position (SEP)^2–4^. Alarmingly, over 75% of global COPD cases^5^ and 90% of asthma cases^6^ occur in low-and middle-income countries (LMICs). These regions often face under-resourced healthcare systems, lack of comprehensive policy and practical local guidelines for CLD diagnosis and management, contributing to low rates of diagnosis and treatment^7,8^. Despite this high burden, the minority of global CLD research funding is concentrated in LMICs, often focusing predominantly on smoking, and less on other critical risk factors like household air pollution (HAP) due to cooking and heating behaviors ^9^.

To design effective interventions targeting these risk factors, it is essential to understand the beliefs, perceptions and behaviors regarding CLD in LMICs. Researchers from the FRESHAIR project^10^, an implementation research project that aimed to improve lung health in low-resource settings, emphasized the necessity of understanding local needs, engaging key stakeholders, and ensuring adequate access to knowledge^11^. In many LMICs, disease beliefs, perceptions, and behaviors are shaped by deeply rooted traditions and cultural norms. For example, smoking may symbolize masculinity, coal-burning rituals is practiced in post-partum care (e.g. Vietnam), and CLD like symptoms might be attributed to witchcrafts or spirits (e.g. Kyrgyzstan)^12^. These indicate limited awareness and increased vulnerability to CLD risk factors among local populations. In Bangladesh, CLD patients rarely know the disease and causes ^13^. Similarly, in rural Uganda, a lack of awareness and health seeking behavior, fear of diagnosis, hindered the adoption of the COPD screening program^14^. These examples underscore the importance of understanding the local beliefs, perceptions and behaviors for developing contextual and successful interventions.

Kyrgyzstan is a relevant and under-researched example within this context. It is a landlocked, mountainous country in central Asia, with 94% of its land above 1000 meters and an average elevation of 2750 meters above sea level^15^. CLD is recognized as one of the main non-communicable diseases in the country^16^, with COPD particularly imposing a substantial clinical and economic burden^17^. Contributing factors include high altitude and extreme climate, widespread smoking, and limited access to clean fuels for cooking and heating^16^ ^18^ ^19^. Highland and lowland populations however differ substantially in risk exposure and lifestyle. Highland communities face harsher weather and, consequently rely more on biomass fuel for cooking and heating, while mountainous terrain also hinders local infrastructure development ^20^. In contrast, lowland areas have milder climates and better access to healthcare, education and infrastructure. Nonetheless, both settings face challenges such as limited healthcare access, shortages of medical supplies and trained staff, and underfunding^21^, hindering disease diagnosis, treatment, and care. Despite the high prevalence of CLD in Kyrgyzstan^17^, few studies explored in depth beliefs, perceptions, and behaviors regarding CLD among local populations.

To achieve the global goal of a 25% reduction in premature mortality from main non-communicable diseases (NCDs), including CLD by 2025, one of the World Health Organization package of essential NCD interventions (PEN) was initiated in Kyrgyzstan in 2015^22^. PEN is a conceptual framework for strengthening equity and efficiency in primary health care in low-resource settings. However, the implementation of PEN in Kyrgyzstan appears to be ineffective in prespecified performance and effectiveness indicators ^22^.

The current study aims to explore the beliefs, perceptions, and behaviors of local community members (CMs) regarding CLD in Kyrgyzstan. Our insights will ultimately provide a better understanding of the local barriers in CLD prevention and care, contributing to the global and sustainable development goals for addressing NCDs. Using a mixed-method study design, we collected data through semi-structured interviews, focus groups, and questionnaires to address the following research questions within the context of Kyrgyzstan:

1. What are the beliefs and perceptions regarding CLD symptoms among CMs in the highlands and lowlands?
2. What are the risk behaviors and help-seeking behaviors of CMs related to CLD in the highlands and lowlands?
3. How do demographics, setting (highlands vs lowlands) and behaviors regarding CLD relate to illness perceptions?
4. What subgroup profiles can be identified based on illness perceptions among CMs?

## Methods

### Study design

This is an exploratory mixed-method study as part of the FRESHAIR project^10^, focusing on the prevention, diagnosis, and treatment of CLD in low-resource settings (see ^10^ for more details). Data were collected in Kyrgyzstan highlands (Naryn) and lowlands (Chui) in May 2016. Qualitative data collection included individual in-depth interviews and focus groups. Quantitative data was collected with questionnaires. All Data collection tools (topic lists and questionnaires) were guided by a theoretical framework that combined three health behavioral models: the Theory of Explanatory Models of Illness^23^, the Theory of Planned Behavior^24^, and the Health Belief Model^25^. Details were published previously^12^.

### Participants and sampling

We included the following participants: 1) CMs aged at least 18 years; 2) Key informants (KIs) with either in-depth knowledge or an overall overview of community beliefs, perceptions and behaviors: e.g., religion or community leaders, traditional healers, pharmacists and teachers. People were excluded if they were not able to participate in the study due to severe physical or mental illness or could not provide formal informed consent.

For the qualitative data collection, purposive sampling was applied to reach a diverse population with sufficient variation in sex, age, educational level and profession. Potential participants who met the inclusion criteria were identified based on snowball sampling and opportunity. Sample size was determined based on data saturation when no new themes or information were emerging from collected data. For the quantitative part, a three-stage sampling approach was applied to randomly select CMs, as published previously^26^. We expected to reach 200 CMs per setting.

### Procedure

Rapid Appraisal approach^27^ was applied to collect qualitative and quantitative data in an effective and iterative way. Researchers immersed themselves into the field and built trust with CMs in advance.

#### Qualitative data

Semi-structured interviews and focus groups were performed by an experienced qualitative researcher and a trained local healthcare researcher for English interpreting. Interviews and focus groups took place in a private place to encourage participants to speak more freely, each lasting approximately 1 hour. Each focus group consisted of 3-6 participants with the same level of hierarchy to avoid hierarchical influence that could inhibit open discussions. Audio was recorded during interviews and focus group. Fieldnotes were recorded when necessary. At the end of each qualitative data collection, a short summary and reflection were discussed with team members.

#### Quantitative data

Hardcopy questionnaires were distributed to CMs by researchers after a pilot test. Additional explanation was provided by researchers in case of any difficulty in understanding questionnaires for participants. Daily meetings after data collection were held for experience exchanges and context-specific adjustments were made according to CMs’ feedback (e.g., rainy/dry season instead of summer/winter)^26^.

### Measures

All measures were described based on the FRESHAIR protocol, details published previously^12^.

#### Interviews and focus groups

Interviews and focus groups were guided by a topic list based on the theoretical framework. After an introduction and obtaining informed consent, demographic information (gender, age, education, profession) was collected. A vignette was used, presenting a case scenario of a relatively young woman, Anna, with CLD signs to avoid potential stigmatization. Questions and probes were used to explore participants’ beliefs and perceptions about CLD, smoking, cooking and heating behaviors, and help-seeking behaviors for CLD. The topic list was tailored to CMs and KIs, and to interviews and focus groups.

#### Questionnaires

##### Demographics

Demographics asked included gender, age in years, education (in years, “In total, how many years have you spent at school and in full-time study?”), and occupation (1=housewife/man, 2=unemployed (able to work), 3=unemployed (unable to work), 4=retired, 5=traditional farming and agricultural service, 6=education or social services, 7=healthcare service, 8=manufacturing, 9=transportation, 10=construction, 11=others). ‘other’ occupations category, was recategorized into unemployed (housewife/man, retired, unemployed) and employed (all other categories).

##### Beliefs and perceptions about CLD

Beliefs and perceptions about CLD were measured with the revised Brief Illness Perception Questionnaire (B-IPQ)^28^. The B-IPQ is a validated questionnaire consisting of nine items (consequence, timeline, personal control, treatment, identity, concern, understanding, emotion, and cause) which can be applied to various diseases, and has good reliability and validity^28^. The B-IPQ employs a 0-to-10 response scale. During pilot testing the original 10 scale was not understood as intended. To ensure internal validity we adapted the response scale into a 4-point scale (1=not at all, 2=slightly, 3=fairly, 4=very well) except item 2 (1=for days, 2=for weeks, 3=for months, 4=for years). The item 9 was originally an open question about causes of CLD. As we expected local CMs were not familiar with CLD causes, we replaced item 9 with 24 potential options and asked them to score whether they think these are causes of CLD with a 5-point Likert scale (from 1=strongly disagree to 5=strongly agree), as was done in FRESHAIR project before^12^. The total B-IPQ score was calculated by summing all 8 items, with item 3, 4, 7 reverse-scored to ensure consistency in interpreting higher scores as reflecting more negative or severe illness perceptions, ranging from 8 to 32.

##### Behaviors regarding CLD

We assessed CMs’ intended help-seeking behavior, smoking, and household risk behaviors such as cooking, heating, and ventilation. Intended help-seeking behavior was assessed by asking “Suppose you had the same condition as Anna, who would you seek for help (partner, friend, traditional healer, general practitioner or primary care doctors, would not seek help etc.)”.

Smoking behavior was assessed by asking “Do you currently smoke any tobacco products, such as cigarettes, cigars or pipes (yes vs no)”, “Do you currently smoke tobacco products daily (yes vs no)” and “Did you smoke daily in the past (yes vs no)”.

For later analysis, smoking was recategorized into no smoking, current smoking and previous smoking.

To quantify the household risk behavior, the household risk behavior score was calculated. Cooking or heating with open or surrounded fire, using risk fuels (e.g., dung, grass, wood, chalk, kerosene), and lack of ventilation were scored 1 for each factor. Considering potential season differences, summer and winter were scored separately. The total score of household risk behavior was then calculated as the sum of these factors, ranging from 0 to 12. Higher scores indicate more frequent household risk behaviors.

### Reflexivity

The research team comprised both local and international researchers from Kyrgyzstan, The Netherlands and China, with diverse backgrounds in respiratory medicine, general practice, psychology, public health, epidemiology, and implementation science. This diversity enriched our interpretations and helped to mitigate bias through team discussions, peer debriefing, and consultation with local stakeholders. Additionally, we are aware of the risks of smoking and HAP, which shaped our interpretations to the data.

### Data analyses

Triangulation was conducted during data analyses. After themes were identified from qualitative data, quantitative findings were compared and complemented to provide more detailed information.

#### Qualitative data

For RQ1 and RQ2, a Framework Method^29^ was used to identify themes emerged from qualitative data. Data were transcribed verbatim and translated into English by the local interpreters who are proficient in Kyrgyz, Russian and English. Transcripts were imported into ATLAS.ti 19.0 for coding and analysis. Each transcript was assigned an study ID. An initial set of open codes were developed inductively and deductively based on 3-5 transcripts with the theoretical framework by two independent researchers. Then these codes were applied to the rest of transcripts. After coding was completed, codes were grouped into categories and then organized into themes and subthemes. All codes and quotes were discussed and reviewed by team members (CS, MK, EM), with discrepancies resolved though consensus. Feedback from Kyrgyz researchers (MM, TS) was incorporated to ensure consistency and validity.

#### Quantitative data

For RQ1 and RQ2, descriptive analyses were conducted by reporting frequencies and percentages for categorical variables, means (M) and standard deviations (SD) for normally distributed continuous variables, and medians and interquartile ranges (IQR) for non-normally distributed variables.

For RQ3, univariable and multivariable linear regression analyses were conducted in SPSS 22.0. Demographics, setting, and behavioral indicators were entered into regression analyses separately as determinants, and total B-IPQ scores as the dependent outcome. All variables with significant associations with total B-IPQ scores (*p*<0.05) were then entered into a multivariable regression model in the same step. For RQ4, latent class analysis (LCA) was applied to B-IPQ scores to identify potential subgroups. The poLCA package^30^ was used in R 4.4.2. A series of LCA models were fit by starting with a one-class model and iteratively adding an additional class. The best model was selected based on the likelihood ratio test (LL), the Bayesian Information Criterion (BIC), Akaike Information Criterion (AIC), relative entropy and parsimony^31^. Lower LL, BIC, AIC indicate better models, and the BIC has been considered as the accurate fit measure. Relative entropy >0.8 indicates a good certainty in classification^32^. We performed LCA by fitting models from 1 to 5 classes with a maximum of 1000 iterations and repeated each analysis 100 times. Default settings were used for other options. After LCA, independent sample t-tests, Chi-square tests and Mann-Whitney U tests were conducted to examine whether class membership was associated with demographics, setting, and behavioral indicators. To further characterize the class profiles, t-tests were used to compare each B-IPQ item scores between classes. We ensure that the assumptions of all analyses were met.

### Ethical considerations

Prior to data collection, an explanation of the study purpose, procedures, potential risks and benefits, and data privacy and security were provided and informed consent hardcopies were obtained from all participants by team members. To mitigate risks of sensitive information disclosure and potential burden of participation, we ensured participation was voluntary and informed participants that they could withdraw at any time. Data could be accessed by the core research team members only and all data were pseudonymized to maintain confidentiality. Ethical approval was acquired from the Leiden University Medical Centre Medical Ethical Committee (P16.063;04/15/2016) in the Netherlands and the National Centre of Cardiology and Internal Medicine in Bishkek Ethics Committee (5;03/03/2016) in Kyrgyzstan.

## Results

### Background characteristics

We included 8 individual interviews, 13 focus groups (n=45), and 420 questionnaires (Appendix pp 1). In the qualitative phase, a total of 53 participants were involved, including 25 (47.2%) males and 28 (52.8%) females, with a diverse age and various occupational backgrounds. Questionnaire participants had a mean age of 47.0 years, including 185 (44.0%) males and 235 (56.0%) females. Most participants were housewives/men (141, 33.6%) or worked in the traditional farming and agriculture sector (110, 26.2%). A total of 40 (9.5%) participants were diagnosed with CLD. More details are shown in Table 1.

**Table 1.** Background characteristics of questionnaire participants.

| Variables | Categories | Highland | Lowland | Total |
| --- | --- | --- | --- | --- |
| Total, n (%) |  | 210, 50.0 | 210, 50.0 | 420, 100.0 |
| Age, mean (SD) | In years | 46.0, 13.6 | 47.9, 13.1 | 47.0, 13.3 |
| Gender, n (%) | Male | 94, 44.8 | 91, 43.3 | 185, 44.0 |
|  | Female | 116, 55.2 | 119, 56.7 | 235, 56.0 |
| Education, median (IQR) | In years | 11.0, 3.0 | 11.0, 3.0 | 11.0, 2.0 |
| Occupation, n (%) | Housewife/man | 69, 32.9 | 72, 34.3 | 141, 33.6 |
|  | Unemployed (able to work) | 7, 3.3 | 14, 6.7 | 21, 5.0 |
|  | Retired | 38, 18.1 | 36, 17.1 | 74, 17.6 |
|  | Traditional farming and agriculture sector | 77, 36.7 | 33, 15.7 | 110, 26.2 |
|  | Education or social service | 15, 7.1 | 20, 9.5 | 35, 8.3 |
|  | Healthcare service | 0, 0 | 7, 3.3 | 7, 1.7 |
|  | Manufacturing, transportation, construction | 0, 0 | 25, 11.9 | 25, 6.0 |
|  | Others<sup>a</sup> | 4, 1.9 | 3, 1.4 | 7, 1.7 |
| Diagnosed with CLD, n (%) | Yes (vs. no) | 31, 14.8 | 9, 4.3 | 40, 9.5 |
| Smoking status, n (%) | No smoking | 151, 71.9 | 139, 66.2 | 290, 69.0 |
|  | Currently smokes | 28, 13.3<sup>b</sup> | 30, 14.3<sup>c</sup> | 58, 13.8 |
|  | Previously smoked | 31, 14.8 | 41, 19.5 | 72, 17.1 |

a: including entrepreneur (n=1), guard (n=4), seller (n=1), student (n=1).
b: male (28, 6.7%), female (0, 0%).
c: male (28, 6.7%), female (2, 0.5%).

### Beliefs, perceptions and behaviors regarding CLD

#### Theme 1: misperceptions about CLD and its causes

##### Misperceiving CLD symptoms as acute, infectious conditions

Most participants demonstrated limited understanding of CLD, frequently misperceiving the symptoms presented in the vignette as a common cold, flu or tuberculosis. For instance, one participant commented *“Maybe, she has got infected from someone, hasn’t she? Maybe flu…If the cough lasts for not more than one month, it might be an ordinary cough” (CM-FG-8-L).* Similarly, a KI noted *“They cough mainly when they have flu. We do also hold the explanatory works on the topics of tuberculosis…If the cough lasts for 2 weeks and this is the sign of the tuberculosis…” (KI-I-11-H)*. Such responses suggest that persistent respiratory symptoms are often perceived as acute or infectious conditions in nature, rather than chronic noncommunicable diseases. This misperception may be reinforced by the “explanatory works”, namely local health education campaigns, which historically placed greater emphasis on infectious diseases than on chronic diseases, as mentioned by this KI.

COPD was not a familiar concept to CMs. When asked what COPD is, one participant responded, *“That’s the first time I have heard this (l*а*ugh)” (CM-FG-5-L).* Similarly, when asked if Anna could have asthma, there were notable misconceptions about its symptomatology and chronic nature. For example, *“Participant: While having asthma, lungs will be squeezed and there will be shortness of breath, but no cough. Interviewer: Do you all agree that it is not asthma? Participant: Yes, we agree (laugh)” (CM-FG-2-H).* Similarly, a KI stated that *“The fact is that there is a shortness of breath, it might progress to complications and become asthma, but in 2-3 years it is hardly possible for it to become asthma.” (KI-I-13-H).* Participants seemed to be more familiar with the term “asthma” than “COPD” but had limited and sometimes inaccurate knowledge about its symptoms and progression. Additionally, participants had difficulty in distinguishing different respiratory illnesses, as illustrated by this KI: *“Interviewer: They can’t distinguish the diseases. Participant: Yes, for all respiratory diseases, they call the illness of the lungs…” (KI-I-13-H).* This reflects a lack of disease differentiation and the tendency to lump all respiratory conditions under a general category.

A few participants also mentioned other conditions such as allergy and bronchitis, although these statements were mostly anecdotal, stemming from relatives or neighbors with similar symptoms and mass media. This suggests their reliance on informal or non-evidence-based source of information, which may lack accuracy and limit their understanding of CLD. For example: *“I have an acquaintance. She retired and coughed all the time from dust and when the flowers and grass were blooming. Allergic.” (CM-FG-6-L)*.

##### Attribution of CLD to cold and smoking

Regarding possible causes of the illness raised in the vignette, most CMs attributed it to cold, referring either to infection or to cold weather in both highland and lowland settings. For example, one mentioned: *“It should be only because of the cold. When you have fever because of the cold and don’t treat yourself, it becomes chronic” (CM-FG-1-L).* The reference to “fever because of the cold” implies they believe it was caused by external microbes, which, if left untreated, could develop into a chronic condition. Another participant, who lived in the highlands, emphasized the role of climatic conditions: *“Here most of the diseases are connected with the highlands, it is cold here. Because of the cold there are health problems with the lungs and other inner organs” (CM-FG-4-H).* This highlights a prevailing belief that cold weather is a direct cause of lung diseases, and even other inner health issues. In addition, some participants acknowledged behavioral factors, such as smoking, as causes of CLD symptoms. For instance, they replied *“This is from the cigarettes, most probably.” (CM-FG-2-H)* and *“Probably, she smokes cigarettes. This can be a reason, or her husband smokes” (CM-FG-8-L).* These statements reflect a partial recognition of smoking— both active and passive smoking— as a cause of CLD, although with uncertainty and limited understanding of the causal relationship.

These qualitative findings were supported by quantitative results. Among 24 potential causes we provided, more than 70% of CMs (strongly) agreed that a germ or virus, kitchen smoke, poor medical care in the past, pollution in the environment, dust, smoking, secondhand smoke, the weather, an allergy, lung infection or tuberculosis in the past, occupational pollution, smoke exposure of mother during pregnancy were causes of CLD, details in Appendix pp 1.

#### Theme 2: Continued smoking despite perceived harms and prevention efforts

##### Smoking is a common behavior shaped by gender norms and role models

Most participants stated that smoking was prevalent in the local community, although they highlighted age and gender differences in smoking prevalence. A village leader in the highlands reflected on this as follows: *“It (smoking prevalence) seems to me 75%…when I look at people standing beside me, there are only a few who do not smoke” (KI-I-13-H)*. A participant in the lowlands estimated that *“around 50% of youngsters smoke, and elderly people do not smoke a lot” (CM-FG-11-L)*. In youngsters, smoking appeared to be related to their paternal practice and peer imitations. Participants noted *“If the father smokes, then the child can also start…In the families where the father does not smoke, it is easier, the child tries not to smoke” (CM-FG-1-L),* and *“there are some in the city, or some of them who study in the city, when they come back, they continue smoking…they have new friends there, who smoke” (CM-FG-4-H).* In the survey, 58 (13.8%) CMs reported to smoke currently and 72 (17.1%) reported previous smoking (Table 1). The use of naswar, a traditional smokeless tobacco, was also prevalent: *“People use naswar more than smoking cigarettes. Naswar gives more pleasure. Men use it more than women, especially the elderly” (CM-FG-5-L).* Male smoking was widely tolerated, whereas female smoking was considered inappropriate. For example, a CM stated that *“the man can be understood… but when women smoke, they are future mothers after all” (CM-FG-5-L).* Another CM explained that *“If girls smoke, it looks a bit strange…but smoking fits them (boys) better than girls” (CM-FG-8-L).* These statements suggested local CMs held different gendered perceptions about smoking, tending to overlook parental roles to men who smoke. One village leader in the highlands added that smoking was linked to idle time during grazing: *“In distant villages, smoking became a reason for inhabitants, because most of the time is free and they get distracted by cigarettes, when they herd their sheep or water the fields, they smoke cigarettes…” (KI-13-I-H)*.

##### Anti-smoking efforts with limited effectiveness and weak enforcement

Anti-smoking efforts were taking place in schools and communities, including brochures, posters and lectures. As a village leader mentioned, “*We gather 8,9,10,11 classes, play and hold the action where we tell about the harm of the smoking and give the rewarding prizes, there we have brochures…do some posters” (KI-FG-2-H).* Another village leader, also a member of the village health committee, remarked *“we read lectures, show movies about the harm of smoking and naswar, show posters, make seminars with all the visual hangouts” (CM-FG-1-L).* Despite these efforts, tobacco control remained weak, especially regarding youth access: *“We strictly prohibit selling cigarettes to children…We just control and threat in words, but they find the ways” (KI-13-I-H).* Notably, CMs were aware of harms of smoking but highlighted the difficulty of quitting. As one participant mentioned *“of course we know that it is harmful, but if you have started, it is hard to give up smoking” (CM-FG-10-L)*.

#### Theme 3: Cooking and heating with risk fuels and limited ventilation

Risk cooking and heating behaviors were noticed, exposing CMs to potential HAP. CMs reported using a kind of large pot, a “kazan”, with open fire, a mix of coal and biomass such as dung and wood for cooking and heating. One participant noted, *“We cook in the kazan outside on the open fire and use biomasses. In winter when it is cold we cook at home…All of us use coal and biomasses” (CM-FG-3-H).* Similarly, another participant shared, *“everybody uses (coals) here” (CM-FG-7-L)*. Although many CMs acknowledged the health risks associated with smoke from coal and biomass, these fuels remained the most commonly used due to their relatively lower cost and ability to retain warmth: *“Coal…keeps the warmth for a long time…it is expensive, but cheaper than electricity” (CM-FG-10-L)*. Ventilation practices were mentioned as partial solutions to reduce smoke exposure, as one participant noted *“when the chimney is clogged, then, the smoke comes out and we clean the chimney” (CM-I-17-H).* Another participant added: *“the smoke comes out, that is why we open windows and doors…We open the doors for 5 minutes (in winter), not so long, while the fire blazes up” (CM-FG-10-L)*. This suggests these ventilation practices were generally limited and dependent on weather conditions. Although electronic oven and gas were seen as cleaner alternatives, their use is constrained by affordability, as noted by CMs in the lowlands: *“We buy gallons of gas and use it only when there is no electricity…It (electricity) is very convenient, but to use it constantly there is big money needed, and we cannot afford them with our salaries” (CM-FG-10-L)*.

The survey demonstrated similar household risk behavior characteristics. As shown in Table 2, many CMs used risk fuels for cooking (summer: 193, 46.0%; winter: 328, 78.1%) and heating (summer: 51, 12.1%; winter: 413, 98.3%) with a noticeable seasonal difference. Access to clean fuels for heating was quite limited (summer: 2, 0.5%; winter: 10, 2.4%). Risk fuel use seemed much more common in the highlands compared to lowlands. Some households had no ventilation when cooking, especially in winter (summer: 3, 0.7%; winter: 83, 19.8%). In the highlands, no ventilation for cooking and heating was common both in summer and winter (Table 2). A sharp increase in reporting no ventilation was noticed in the lowlands, from 0% in summer to 36.2% in winter, suggesting greater sensitivity to seasonal changes here. Additionally, participants had a median household risk behavior score of 5.0 (IQR 1.0) in summer and 7.0 (IQR 0.0) in winter (range: 2-12). This suggests CMs engaged in less frequent household risk behaviors in summer and more frequent in winter. More details are shown in Table 2.

**Table 2.** Household risk behaviors of community members.

| Categories |  | Summer (n, %) |  |  | Winter (n, %) |  |  |
| --- | --- | --- | --- | --- | --- | --- | --- |
|  |  | Highland | Lowland | Total | Highland | Lowland | Total |
| Risk cooking stoves | open/surrounded fire | 25, 11.9 | 7, 3.3 | 32, 7.6 | 1, 0.5 | 0 | 1, 0.2 |
| Cooking fuels | risk fuels<sup>a</sup> | 144, 68.6 | 49, 23.3 | 193, 46.0 | 201, 95.7 | 127, 60.5 | 328, 78.1 |
|  | clean fuels | 121, 57.6 | 179, 85.2 | 300, 71.4 | 18, 8.6 | 153, 72.9 | 171, 40.7 |
| Cooking ventilation | no ventilation | 3, 1.4 | 0 | 3, 0.7 | 7, 3.3 | 76, 36.2 | 83, 19.8 |
| Risk heaters* | open fire/surrounded fire | 0 | 0 | 0 | 0 | 0 | 0 |
| Heating fuels | risk fuels | 51, 24.3 | 0 | 51, 12.1 | 210, 100.0 | 203, 96.7 | 413, 98.3 |
|  | clean fuels | 2, 1.0 | 0 | 2, 0.5 | 0 | 10, 2.4 | 10, 2.4 |
| Heating ventilation* | no ventilation | 0 | 6, 2.9 | 6, 1.4 | 0 | 6, 2.9 | 6, 1.4 |

\*: no difference between summer and winter.
a: solid fuels (e.g., coal, wood, grass, dung) and kerosene.

#### Theme 4: Delayed help-seeking

##### Traditional remedies for early respiratory symptoms

In both the highlands and lowlands, traditional remedies were frequently mentioned as a way of relieving early respiratory symptoms, such as cough and sore throat, by applying drinks or food mixed with animal fat, meat, milk, and herbs. For example, one participant mentioned: *“The fat of sheep is applied to the throat. Feet are steamed with the mustard flour. Mustard poultice is applied. And for the sore throat, the fat of badger helps, it is necessary to eat it” (CM-FG-5-L).* This demonstrates the local people tended to treat all respiratory symptoms as ailments and the effectiveness of traditional remedies was widely acknowledged.

##### Reliance on self-medication and pharmacies over formal care

If symptoms persisted, people were more likely to buy medications at the pharmacy. One participant described: *“they all do self-treatment, only when the case is very hard, they turn to help…I haven’t seen them to go to the hospital in a large scale” (CM-FG-7-L)*. This reflects a common pattern of delayed help-seeking, where CMs tended to seek professional medical care only in a severe or advanced stage of disease. When asked what medications they took, one participant responded: *“Antibiotics, antiviral…it is easy (to buy antibiotics), if there are no such medications here, we go to the nearest villages and buy there” (CM-FG-4-H).* This implies antibiotics use was quite common, and they were easily bought in local pharmacies. A key reason for choosing self-medication over formal care was related to accessibility and affordability. For example, some participants noted that medical facilities were far away “*it is 25 km to the nearest drugstore” (CM-FG-11-L)* and hospital visits were costly “*financial problems…there are people, who do not want intentionally” (CM-FG-7-L)*. In addition, a CM in the lowlands reported there was a lack of qualified HCPs that can provide corresponding treatment: *“we do not have the doctor; if we had, we would go to consult him…At our medical post our nurses are not allowed even to use drip bulb” (CM-FG-11-L).* In the highlands, it seemed much harder to consult a doctor or paramedic due to the long distance to medical facilities and transportation problems: *“there is no connection, no gasoline, no car. But in summer it is easier. We ride to the border post on horses and get there by car” (CM-FG-2-H).* These statements suggest that distance to medical facilities and financial burden act as a barrier to access timely and basic healthcare services, especially in remote areas with infrastructure issues.

##### Healthcare preference shaped by trust, cost and cultural background

When formal care was sought, CMs expressed a greater trust in private hospitals compared to state hospitals, despite higher costs. One participant pointed out the different quality of care between private and state hospitals: *“At state hospital, the doctors… well, I think, they do not treat well…they can make a wrong diagnosis…But the private hospitals are better…those who can afford this, they turn to private ones. And those who cannot afford they turn to the state hospital…many will go to the state…” (KI-FG-1-L).* This implies CMs with high income could access better healthcare service while those with lower income were left with lower quality of care. Notably, help-seeking behavior also appeared to be linked to ethnic and cultural backgrounds, as a participant with a Kyrgyz ethnical background mentioned: *“In Bishkek I have Russian neighbors, if they have a small problem with their health, they immediately go to see the doctor…We are nomad nation. We’re used to herd cattle and didn’t know any doctors…” (CM-FG-11-L).* This contrast between Russians and Kyrgyz ethnicities was mentioned by several other participants.

According to the survey, however, most CMs would seek help from general practitioner or primary care doctors (highland: 201, 95.7%; lowland: 201, 95.7%) when encountering Anna’s condition. Turning to local nurses was also a preferred option (highland: 130, 61.9%; lowland: 94, 44.8%). Only 7 (1.7%) CMs in the lowlands would not seek help. This reveals a notable discrepancy between participants’ intended and actual help-seeking behaviors.

### Explaining B-IPQ scores

CMs obtained a mean B-IPQ score of 23.4 (SD 2.1), with 23.1 (SD 2.2) in the highlands and 23.7 (SD 1.9) in the lowlands. Univariable and multivariable linear regression results for B-IPQ scores are shown in Table 3. In the univariable analyses, longer duration of education, unemployment, and living in the lowlands were significantly associated with higher B-IPQ scores (*p*<0.05). All associations were still significant in the multivariable model (*p*<0.05).

**Table 3.** Univariable and multivariable regression analyses for B-IPQ scores.

| Variables | Categories | B-IPQ<sup>a</sup><br>scores (M,<br>SD) | Univariable regression |  |  | Multivariable regression |  |  |
| --- | --- | --- | --- | --- | --- | --- | --- | --- |
| | | | *b* (95% CI) | $\beta$ | *p* | *b* (95% CI) | $\beta$ | *p* |
| Age in years |  | 23.4, 2.1 | -0.00 (-0.02, .01) | -0.01 | .77 |  |  |  |
| Gender | Male | 23.2, 2.1 | Reference |  |  |  |  |  |
|  | Female | 23.5, 2.0 | 0.38 (-0.02, 0.78) | 0.09 | .06 |  |  |  |
| Education in years |  |  | 0.11 (0.02, 0.20) | 0.11 | .021* | 0.10 (0.00, 0.19) | 0.10 | .045* |
| Occupation | Unemployed | 23.6, 1.9 | Reference |  |  | Reference |  |  |
|  | Employed | 23.1, 2.3 | -0.49 (-0.89, -0.09) | -0.12 | .016* | -0.48 (-0.87, -0.08) | -0.11 | .018* |
| Setting | Highland | 23.1, 2.2 | Reference |  |  | Reference |  |  |
|  | Lowland | 23.7, 1.9 | 0.59 (0.20, 0.99) | 0.14 | <.01* | 0.52 (0.13, 0.92) | 0.13 | .01* |
| Smoking status | No smoking |  | Reference |  |  |  |  |  |
|  |  | 23.4, 2.1 |  |  |  |  |  |  |
|  | Currently smokes | 23.5, 1.9 | 0.10 (-0.49, 0.69) | 0.02 | .73 |  |  |  |
|  | Previously smoked | 23.3, 2.1 | -0.13 (-0.67, 0.41) | -0.02 | .64 |  |  |  |
| Household risk behavior scores in summer |  |  | -0.02 (-0.18, 0.15) | -0.01 | .82 |  |  |  |
| Household risk behavior scores in winter |  |  | -0.01 (-0.33, 0.31) | -0.00 | .96 |  |  |  |

a: brief Illness Perception Questionnaire.
\*: $p<0.05$ .

### LCA model

The LCA model characteristics are shown in Table 4. The model with two classes (model 2) was considered as the best fitting model, with the lowest BIC and a high relative entropy value, indicating high certainty of classification. Figure 1 depicts the conditional item response probabilities for B-IPQ items in the two classes, with 49.3% of population share in class 1 and 50.7% in class 2. Overall, CMs in class 1 appeared to have a more negative perception about the CLD consequence, symptoms, concern and emotion, whereas those in class 2 tended to perceive CLD timeline, treatment and understanding more negatively (Figure 1). T-tests showed significant differences in all B-IPQ items between the two classes (*p*<0.05) except for control (*p*=0.67). Class comparison of participant characteristics was shown in Appendix pp 2. CMs in the highlands were more likely to belong to class 1 (χ^2^=5.95, *p*=0.01). No other factors were significantly related to class membership (Appendix pp 2).

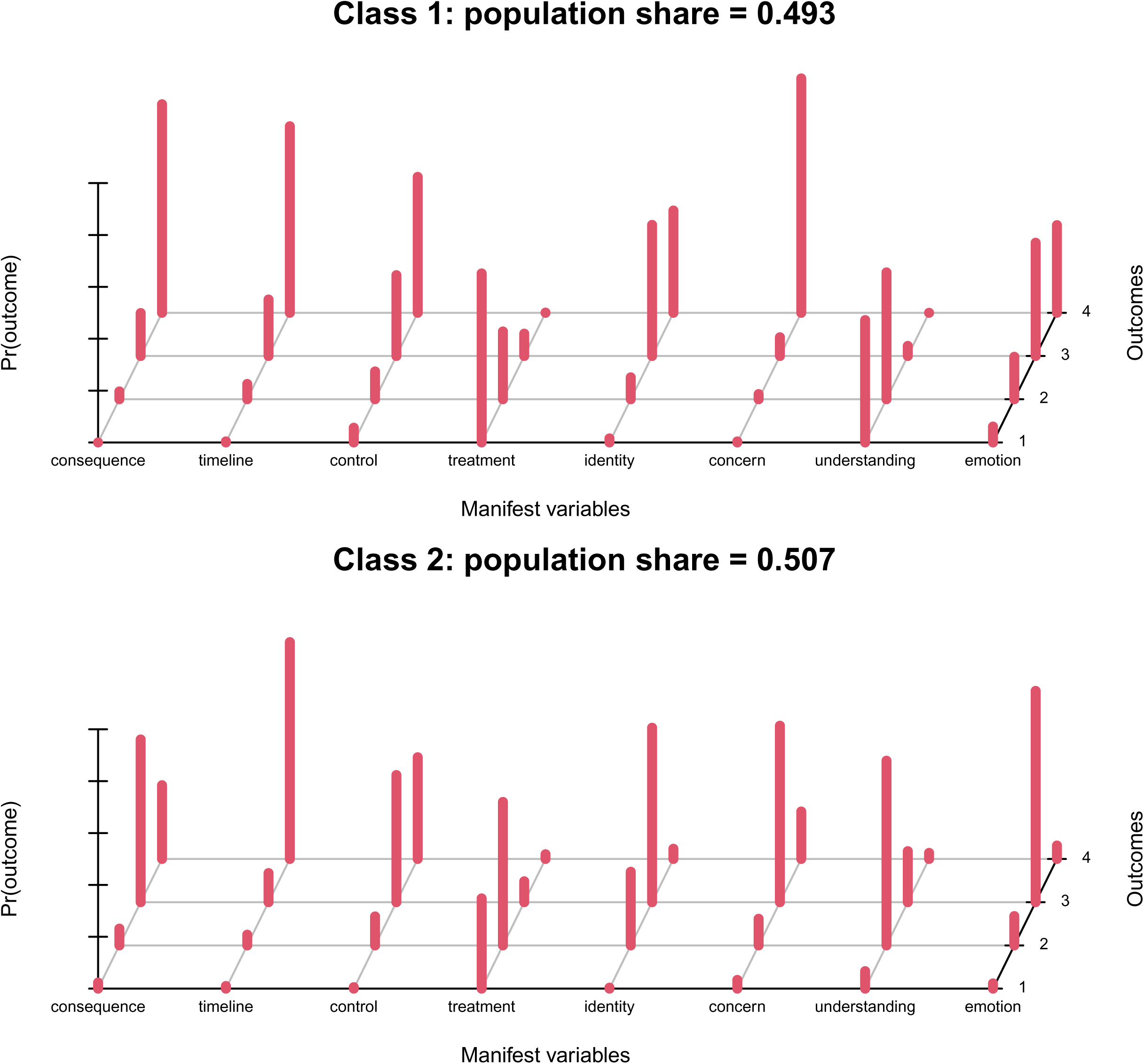

**Table 4.** Latent class analysis model characteristics for community members.

| Classes | LL<sup>a</sup> | BIC<sup>b</sup> | AIC<sup>c</sup> | Parameters | Entropy | Relative entropy | Class membership<sup>d</sup> |
| --- | --- | --- | --- | --- | --- | --- | --- |
| 1 | -3164.715 | 6474.396 | 6377.429 | 24 | 7.535 | 1.00 | 100% |
| 2 | -3003.726 | 6303.424 | 6105.452 | 49 | 7.162 | 0.975 | 1: 49% |
|  |  |  |  |  |  |  | 2: 51% |
| 3 | -2946.602 | 6340.183 | 6041.204 | 74 | 7.026 | 0.985 | 1: 33% |
|  |  |  |  |  |  |  | 2: 52% |
|  |  |  |  |  |  |  | 3: 15% |
| 4 | -2913.16 | 6424.304 | 6024.319 | 99 | 6.945 | 0.949 | 1: 44% |
|  |  |  |  |  |  |  | 2: 28% |
|  |  |  |  |  |  |  | 3: 9% |
|  |  |  |  |  |  |  | 4: 19% |
| 5 | -2894.947 | 6538.886 | 6037.894 | 124 | 6.887 | 0.965 | 1: 23% |
|  |  |  |  |  |  |  | 2: 24% |
|  |  |  |  |  |  |  | 3: 8% |
|  |  |  |  |  |  |  | 4: 37% |
|  |  |  |  |  |  |  | 5: 7% |

a: Maximum log-likelihood.
b: Bayesian Information Criterion.
c: Akaike Information Criterion.
d: Values represent estimated population class shares.

## Discussion

### Main findings

Building on previous findings of the FRESHAIR project, this mixed-method study offers novel and more detailed insights into beliefs, perceptions and behaviors of community members (CMs) regarding CLD in the highlands and lowlands Kyrgyzstan. These findings can provide contextual evidence to inform future implementation research in other low-resource settings or LMICs. In our study, we found that misperceptions and misbeliefs existed regarding CLD and its causes, along with prevalent smoking, cooking and heating with risk fuels and limited ventilation, delayed help-seeking behaviors. These beliefs, perceptions, and behaviors were shaped by socioeconomic, cultural and policy-related factors. Additionally, our results showed that CMs with longer duration of education, being unemployed, and living in the lowlands tended to have more negative CLD beliefs and perceptions. LCA further identified two subgroups with different B-IPQ profiles: 49% of CMs in class 1 had more negative perceptions in the B-IPQ consequence, symptoms, concern and emotion, while 51% in class 2 perceived more negatively in the B-IPQ timeline, treatment and understanding. Highlanders were more likely to be in class 1 while lowlanders in class 2.

### Comparison with prior research

The misperceptions and misbeliefs about CLD indicate a limited understanding of CLD among CMs in Kyrgyzstan. This finding aligns with the main results of the FRESHAIR project conducted in other low-resource settings, including Uganda, Greece, Vietnam and a Roma camp^12^, where CLD symptoms were also perceived as infectious conditions. Comparable findings were reported in a qualitative study^33^ and a mixed-method study^34^ in China, both of which highlighted gaps in awareness and understanding of CLD. In our study, the term COPD was unfamiliar to many CMs, and knowledge about asthma symptoms was also inaccurate, suggesting the absence of the targeted health education and promotion on CLD. Local health campaigns in Kyrgyzstan seemed to place greater emphasis on infectious diseases such as tuberculosis, than on chronic conditions like CLD. As a result, symptoms associated with CLD may be overlooked or misinterpreted through the lens of infectious diseases. To some extent, this reflects the existing public health priorities on infectious diseases and insufficient effectiveness of health educational campaigns promoting knowledge of CLD^35^. Contributing CLD condition to cold and weather might be because of the distinctive geography and climate in Kyrgyzstan, as long-term residence at high altitude and low temperature exposure both can affect pulmonary function^36^, producing symptoms similar to CLD. This may lead local people to normalize coughing and shortness of breath, therefore obscuring the recognition of CLD.

In our study, two latent subgroups were identified with different B-IPQ profiles. Previous studies using B-IPQ in CLD have reported similar domain-specific patterns. For example, Dutch COPD patients had better health-related quality of life when experiencing fewer symptoms, less consequences, fewer emotional impacts and having a greater understanding of the disease^37^. In China, Liu et al.^38^ reported inhaler adherence is also different in COPD patients across different levels of B-IPQ control, treatment, understanding emotions. This suggests that differences in B-IPQ scores between subgroups can be useful for designing tailored interventions in the future.

Our results suggest that socio-economic position (SEP) — including duration of education, employment status, and income —is associated with CMs’ beliefs, perceptions and behaviors regarding CLD. Lower SEP is typically associated with more negative and severe illness perceptions in high-income countries (HICs)^39^ ^40^, but we found longer education duration was associated with more negative perceptions. This illustrates a different role of SEP between HICs and LMICs. A potential explanation might be that individuals with higher education are more exposed to health information^41^, therefore have a better understanding of the chronic and irreversible nature of CLD, raising greater concerns and perceived severity. In our study, unemployed individuals perceived CLD more negatively partly due to their vulnerability and lack of coping strategies, along with more emotional impact such as depression and anxiety^42^. The prevalent smoking among nomadic pastoralists during herding may also be explained by their lower SEP, including limited access to education ^43^ and chronic poverty in remote areas^44^. Moreover, income or financial constraint emerged as a recurring factor that shaped CLD-related behaviors in our study, including cooking and heating with risk fuels, and help-seeking. For instance, participants preferred to use biomass fuels mainly because of the high cost of clean energy, despite having access to electricity. Another study also pointed out additional financial barriers to the implementation of renewable energy supply in Kyrgyzstan, such as the uninsulated and aged building stock^45^. In terms of help-seeking, lower income limits the ability to seek timely and high-quality medical care. This finding aligns with a cross-sectional study from Brazil, which showed an evident disparity in the access to and quality of care services across socio-economic groups^46^.

Cultural factors, including gender norms, ethnic identity and practice of traditional remedies, shaped people’s beliefs, perceptions and behaviors regarding CLD among CMs. Culturally, CMs perceived male smoking as normal while female smoking was stigmatized. They believed that mothers played a larger role during early childhood whereas fathers appeared to become more prominent role models for smoking when kids grew older. The traditional gender norms, mirrors findings from Southeast Asia, where smoking is commonly associated with masculinities^12^ ^47^. The difference in help-seeking behaviors also underscores the cultural context. For example, Kyrgyz ethnicity with nomadic background appeared less likely to seek medical care compared to ethnic Russians. Similarly, another study reported that African American and Hispanic participants were more likely to receive mental health care for multiple sclerosis compared to white and Non-Hispanic participants^48^. The traditional remedies for respiratory health in Kyrgyzstan, which have also been used for centuries in Asia^49^, Africa^50^ and Eastern Europe^51^, underscore the need for culturally sensitive health education. Given that the efficacy and safety of these remedies usually lack scientific validation^50^ and evidence-based guidelines^52^, it is necessary to integrate rigorously evaluated traditional knowledge into public health strategies.

Our findings also indicate policy shortcomings in CLD prevention and care. The prevalence of smoking, especially among men and adolescents, points to the need to accelerate the local enforcement of tobacco control policies and sales regulations, as supported by the latest report on the tobacco epidemic in Kyrgyzstan^53^ and a global implementation research targeting adolescents’ tobacco exposure in disadvantaged populations^54^. Regarding the clean energy adoption, Kedar et al. described multiple challenges in the local context, such as affordability, limited availability, old infrastructure, and heat loss^45^. Knowledge and beliefs about the innovation, external policy and incentives also matter, according to an umbrella review^55^. These highlight the importance of policies aimed at improving energy infrastructure and promoting sustainable energy transitions. Furthermore, the easy accessibility of antibiotics without prescription reflects broader weakness of pharmaceutical regulations, which may contribute to inappropriate use and antimicrobial resistance^56^. Reported barriers to healthcare, such as long travel distance to healthcare facilities, shortage of HCPs and disparities in care quality between state and private hospitals, reflect underlying health inequalities. These inequalities exist both between and within HICs and LMICs, and addressing them requires organized and systematic health planning and policy efforts^57^.

### Implications and Recommendations

The misperceptions about CLD underscore a need for targeted educational interventions in practice. Public health campaigns should focus on increasing awareness of CLD, especially its chronic nature, multiple causes, such as smoking and HAP, and early warning signs. To improve the effectiveness of health campaigns, joint planning and implementation, sharing of information systems, and demand creation and community-based engagement strategies among stakeholders were recommended^58^. The current B-IPQ profiles between two subgroups can help inform the design of context-specific implementation strategies tailored to different populations. To effectively change CM’s risk behaviors and promote actions, behavioral change techniques should be incorporated in health interventions^59^. Given the existing use of local educational tools like posters, brochures and lectures, we recommend the integration of digital technologies into health education efforts as digital health education demonstrated a promising feasibility, usability and user satisfaction to improve the awareness^60^. Involving the community and relevant stakeholders to co-create educational materials with plain language and illustrations also has been proven feasible and effective in LMICs^61^.

For policymakers, it is essential to implement gender-sensitive, community-supported smoking cessation programs^62^ and to strengthen tobacco sales regulations. Improving energy infrastructure, promoting affordable and sustainable clean energy should also be prioritized, aligning with the Sustainable Development Goals^63^. To reduce the health inequality stemming from limited healthcare affordability and accessibility, coordinated efforts are required, such as expanding healthcare services, improving healthcare service delivery, training and retaining qualified HCPs in remote areas. For future research, we recommend further evaluation studies to assess the effectiveness of interventions on CLD awareness and behavioral change. More studies are also warranted on the diagnosis, management and self-management of CLD in primary care in Kyrgyzstan and other LMICs^64^.

### Limitations and strengths

This study has several limitations. First, its cross-sectional design limits causal interpretations. Second, as data were collected in 2016, the findings may have limited relevance to the current infrastructural and systemic context. Third, the findings should be extrapolated with caution due to the sampling methods, and the qualitative findings are not generalizable by nature. Nevertheless, this study provides nuanced and context-specific insights into beliefs, perceptions and behaviors of CM regarding CLD in Kyrgyzstan. The combination of qualitative and quantitative methods complemented each other, enabling a more comprehensive and in-depth understanding of the barriers in primary care in the local context and informing future implementation research. Moreover, the use of LCA identified distinct subgroups based on B-IPQ scores, which cannot be captured by regression analyses alone.

## Conclusion

Misbeliefs and misperceptions in chronic lung diseases existed among community members in Kyrgyzstan, alongside prevalent smoking, household risk behaviors, and a pattern of delayed help-seeking. These beliefs, perceptions, and behaviors were shaped by socioeconomic, cultural and policy-related factors. Community members with longer duration of education, being unemployed and living in the lowlands held more negative disease perceptions. There were two subgroups with different B-IPQ profiles: class one with more negative perceptions in the consequence, symptoms, concern and emotion, while class two perceived more negatively in the timeline, treatment and understanding, which can be used for tailored interventions in different populations. These insights provide context-specific evidence to inform future implementation research aimed at improving CLD awareness, promoting behavioral change and strengthening primary care in Kyrgyzstan and potentially other low- and middle-income countries.

## Data Availability

All data produced in the present study are available upon reasonable request to the authors

## Abbreviations

AIC: Akaike Information Criterion
BIC: Bayesian Information Criterion
CLD: chronic lung disease
CM: community member
COPD: chronic obstructive pulmonary disease
FRESHAIR: Free Respiratory Evaluation and Smoke exposure reduction by primary Health cAre Integrated gRoups
HCP: healthcare professional
HIC: high-income country
KI: key informant
LCA: latent class analysis
LL: likelihood ratio test
LMIC: low- and middle-income country
NCD: non-communicable disease
SEP: socioeconomic position

## Availability of data and materials

The datasets collected and analyzed in the current study are available from the corresponding author on reasonable request.

## Authors’ contributions

In this manuscript, CS contributed to the study design, data analysis, and manuscript writing. EM, MK, NC supervised the study and contributed to the manuscript revision and editing. EAB, CP, RMK, MM, TS contributed to the data collection and manuscript revision and editing. All authors reviewed and approved the final manuscript.

## Acknowledgement

We extend our sincere gratitude to all participants for their contributions to this study, and to the ATLAS.ti Support Team for data recovery. We also acknowledge the researchers and collaborators involved in the FRESHAIR project in Kyrgyzstan, whose efforts laid the foundation for this work.

