## Supplementary material for "Beliefs, Perceptions, and Behaviors Regarding Chronic Lung Disease in Kyrgyzstan: A Mixed-method FRESHAIR Study"

Number of filed activities in quantitative and qualitative data collection

|  | Highland | | Lowland | | Total |
| --- | --- | --- | --- | --- | --- |
|  | CMs | KIs | CMs | KIs | Total |
| Interviews | 2 | 4 | - | 2 | 8 |
| Focus groups | 3 | - | 8 | 2 | 13 |
| Questionnaires | 210 | - | 210 | - | 420 |

Perceived causes checked by community members

| Causes | Highland (n, %)  (N=210) | Lowland (n, %)  (N=210) |
| --- | --- | --- |
|  | (strongly) agree | (strongly) agree |
| Stress or worry | 154, 73.3 | 130, 61.9 |
| It runs in the family | 158, 75.2 | 158, 75.2 |
| A germ or virus | 177, 84.3 | 190, 90.5 |
| Kitchen smoke | 149, 71.0 | 132, 62.9 |
| Diet or eating habits | 150, 71.4 | 124, 59.0 |
| Chance or bad luck | 64, 30.5 | 53, 25.2 |
| Poor medical care in the past | 145, 69.0 | 169, 80.5 |
| Pollution in the environment | 188, 89.5 | 193, 91.9 |
| Family problems or worries | 125, 59.5 | 120, 57.1 |
| Overwork | 105, 50.0 | 98, 46.7 |
| Ageing | 125, 59.5 | 130, 61.9 |
| Dust | 185, 88.1 | 180, 85.7 |
| Alcohol | 139, 66.2 | 141, 67.1 |
| Smoking | 206, 98.1 | 205, 97.6 |
| Second hand smoke | 189, 90.0 | 191, 91.0 |
| Witchcraft | 70, 33.3 | 74, 35.2 |
| The weather | 164, 78.1 | 174, 82.9 |
| Spirits | 41, 19.5 | 57, 27.1 |
| Brought from other regions | 139, 66.2 | 165, 78.6 |
| An allergy | 166, 79.0 | 160, 76.2 |
| Lung infection or tuberculosis in the past | 178, 84.8 | 199, 94.8 |
| Occupational pollution | 198, 94.3 | 196, 93.3 |
| Smoke exposure of mother during pregnancy | 174, 82.9 | 193, 91.9 |
| Exercise | 51, 24.3 | 22, 10.5 |

Class comparison of participant characteristics after LCA

| Variable | Categories | Class 1 (n=212) | Class 2 (n=208) | Statistics |
| --- | --- | --- | --- | --- |
| Gender, n (%) | Male | 83, 39.2 | 102, 49.0 |  |
|  | Female | 129, 60.8 | 106, 51.0 | χ^2^(1)=3.77, *p*=0.052 |
| Age, mean (SD) |  | 46.7, 13.4 | 47.3, 13.2 | t (418)=-0.50, *p* =0.62 |
| Education in years, median (IQR) |  | 11.0, 2.0 | 11.0, 1.3 | W=23917, *p*=0.12 |
| Occupation, n (%) | Unemployed | 119, 56.1 | 118, 56.7 |  |
|  | Employed | 93, 43.9 | 90, 43.2 | χ^2^(1)=0.00, *p*=0.98 |
| Setting, n (%) | Highland | 119, 56.1 | 91, 43.8 |  |
|  | Lowland | 93, 43.9 | 117, 56.2 | χ^2^(1)=5.95, *p*=0.01* |
| Smoking status, n (%) | No smoking | 153, 72.2 | 137, 65.9 |  |
|  | Currently smokes | 25, 11.8 | 33, 15.9 |  |
|  | Previously smoked | 34, 16.0 | 38, 18.3 | χ^2^(2)=2.17, *p*=0.34 |
| Household risk behavior scores in summer, median (IQR) |  | 6.0, 1.0 | 5.0, 1.0 | W=23822, *p*=0.12 |
| Household risk behavior scores in winter, median (IQR) |  | 7.2, 0.7 | 7.1, 0.5 | W=22949, *p*=0.20 |
| *: *p* <0.05. | | | | |
